# Early Real-world Implant Experience with a Helix-fixation Ventricular Leadless Pacemaker

**DOI:** 10.1101/2023.10.13.23297030

**Authors:** Devi G. Nair, Derek V. Exner, Vivek Y. Reddy, Nima Badie, David Ligon, Marc A. Miller, Bridget Lee, Brandon Doty, Athanasios Thomaides, Zayd Eldadah, Malick Islam, Cyrus Hadadi

## Abstract

**Background:** Roughly 1 in 6 patients receiving conventional transvenous pacemaker systems experience significant complications within 1 year of implant, mainly due to the transvenous lead and subcutaneous pocket. A new helix-fixation single-chamber ventricular leadless pacemaker (LP) system capable of pre-deployment exploratory electrical mapping is commercially available. Such an LP may mitigate complications while streamlining the implantation.

**Objectives:** Evaluate the initial real-world implant experience of the helix-fixation LP following its commercial release.

**Methods:** In patients indicated for single-chamber right ventricular pacing, helix-fixation Aveir VR LPs (Abbott, Abbott Park, IL) were implanted using the dedicated loading tool, introducer, and delivery catheter. Implant procedural characteristics, electrical parameters, and any 30-day procedure-related adverse events of consecutive implant attempts were retrospectively evaluated.

**Results:** A total of 167 patients with Class I indication for permanent pacing received implants in 4 North American centers (57% male, 70 years old). Pre-fixation electrical mapping of potential sites allowed repositioning to be avoided in 95.7% of patients. Median [interquartile range] LP procedure and fluoroscopy durations were 25.5 min [20.0, 35.0] and 5.7 min [4.0, 9.2], respectively. Pacing capture threshold, sensed R-wave amplitude, and impedance were 0.8 V [0.5, 1.3], 9.0 mV [6.0, 12.0], and 705 Ω [550, 910], respectively. Implantation was successful in 98.8% of patients, with 98.2% free from acute adverse events.

**Conclusion:** The initial, real-world experience of the helix-fixation ventricular leadless pacemaker demonstrated safe and efficient implantation with minimal repositioning, viable electrical metrics, and limited acute complications.

## Introduction

Despite nearly 70 years of research and clinical experience with traditional pacemakers and transvenous leads, roughly 1 in 6 patients will experience complications within the first year post-implant.^1–3^ Common pacemaker complications are known to easily double the cost of an otherwise uncomplicated procedure.^1, 4^ The majority of these complications – including lead dislodgement, lead fracture, pocket infection, valve regurgitation, hematoma, and pneumothorax – were directly attributed to the transvenous lead and subcutaneous pulse generator pocket.^1–3^ The leadless pacemaker (LP) is a miniaturized alternative to the transvenous pacemaker and is housed entirely within the target chamber. Without a need for transvenous leads or a generator pocket, LPs may mitigate the associated complications^5^ while streamlining the implant procedure and accelerating patient recovery^6^.

The Aveir VR leadless pacemaker (Abbott, Abbott Park, IL) is a commercially available, single-chamber ventricular LP. Features unique to this LP include a helix-based fixation mechanism, pre-deployment exploratory electrical mapping capability, and distinct catheters designed specifically for streamlined delivery and retrieval.^7^ This novel LP system has yet to be systematically evaluated in a real-world clinical setting following its commercial release in the United States. This study reports the initial, real-world implant experience of the LP in four North American Centers.

## Methods

### Study Design

This retrospective study was performed according to the principles outlined in the Declaration of Helsinki and the Good Clinical Practice guidelines of the European Commission. Patients at least 18 years of age who met standard criteria for permanent pacing, and in whom the helix-fixation LP was prescribed were included in the analysis. Records from consecutive patients undergoing implantation at the four participating centers were retrospectively reviewed. All patients provided written informed consent. The LP device, implant procedure, and procedural analyses are described below.

### Device Description

The Aveir VR leadless pacemaker (Abbott, Abbott Park, IL) contains a pulse-generator, battery, helix-fixation mechanism, docking mechanism, and pacing/sensing electrodes designed to provide pacing in the RV, as shown in **Figure 1**. At the distal end, the LP employs an outer helix as the primary fixation mechanism, and a central, inner dome-shaped electrode as the cathode. The proximal end of the LP case serves as the LP anode, separated from the cathode by parylene insulation coating. It also features a docking button that facilitates coupling and torque transfer interactions with the delivery and retrieval catheters.

**Figure 1.**
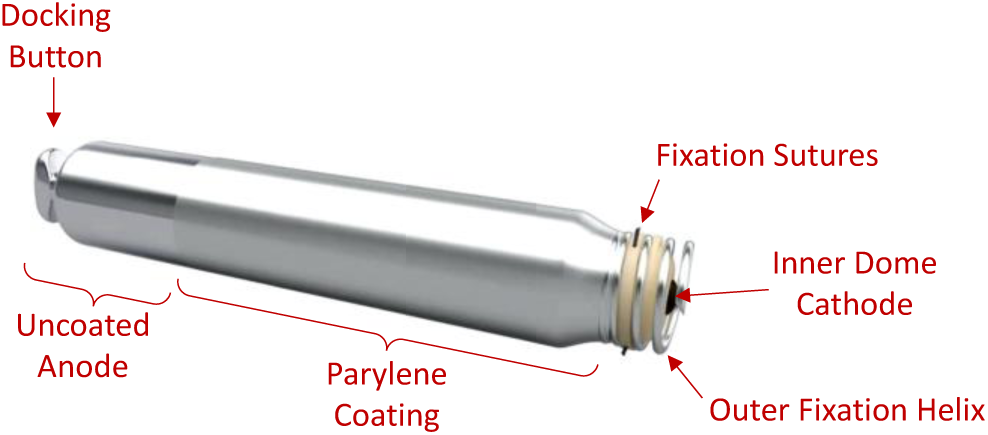
Helix-fixation ventricular leadless pacemaker. The distal outer helix facilitates torque-driven fixation and retrieval; fixation sutures enhance implant stability. The proximal docking button facilitates contact by the delivery or retrieval catheter. Electrical stimulation and wireless communication are achieved via the uncoated case on the proximal end (anode) and inner dome electrode on the distal end (cathode), isolated electrically by parylene insulation coating on the device case.

### Implant Procedure

Each LP device was implanted per standard Instructions for Use (IFU). The 25 Fr inner diameter Aveir Introducer (Abbott) was first placed in the right or left femoral vein via standard percutaneous access. The catheter system includes a steerable delivery catheter, integrated guiding catheter with protective sleeve, and hemostasis valve bypass tool to dilate the introducer sheath hemostasis valve. The LP, pre-packaged in the Aveir Loading Tool (Abbott), was tethered to the Aveir Delivery Catheter (Abbott) and pulled in to fully dock with the delivery catheter docking cap, and the protective sleeve was advanced over the LP. The catheter and LP were advanced through the existing introducer into the femoral vein and fluoroscopically guided through the peripheral vasculature to the RV chamber. **Figure 2** provides fluoroscopic images illustrating an LP delivery example. Intracardiac ultrasound was used as per operator preference.

**Figure 2.**
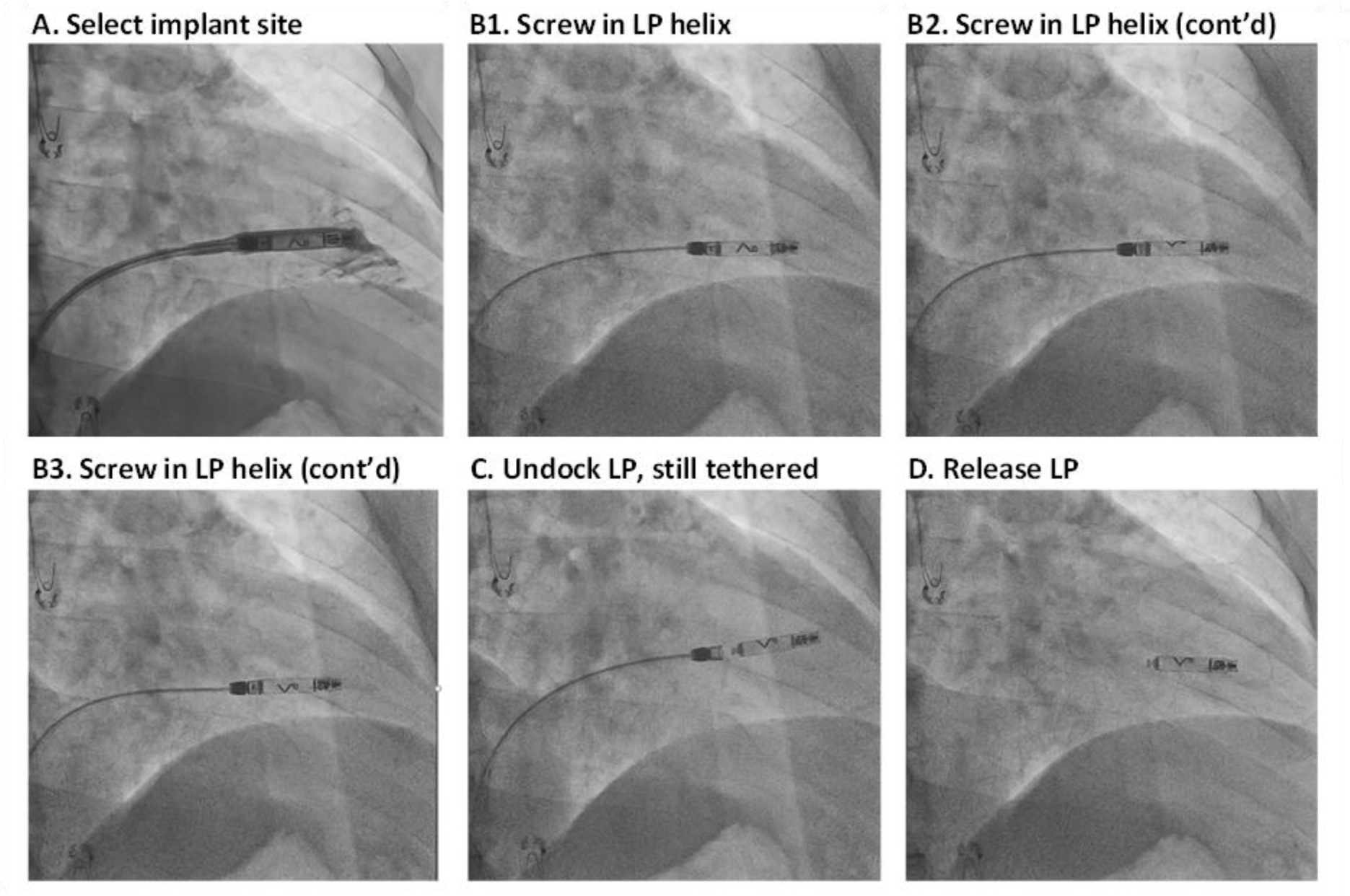
Fluoroscopic images demonstrate an example LP delivery from a right anterior oblique view. (A) Implant site is selected using contrast agent with the protective sheath still in place. (B1, B2, B3) With acceptable electrical metrics, the LP helix is screwed into the myocardium, as visualized by the directional chevron marker rotating clockwise from behind the LP to in front of the LP. (C) LP is undocked from the delivery catheter cap, revealing the LP docking button, yet is still tethered for potential re-docking. (D) With acceptable electrical metrics verified, the LP is released from the delivery catheter tethers.

Once the RV was fluoroscopically surveyed using contrast injection and a potential implant site was selected (**Figure 2**A), the protective sleeve was retracted to expose the LP, and LP-myocardial electrical contact was evaluated by using the Merlin Patient Care System (i.e., programmer; Abbott). Briefly, the Programmer was connected to the Aveir Link Module (Abbott), which served as an interface to electrocardiogram (ECG) patch electrodes on the patient’s torso and provided bi-directional telemetry communication with the LP. The pacing capture threshold (PCT), sensed R-wave amplitude, pacing impedance, and current of injury EGM (COI) electrograms were collected prior to deployment, with the LP still fully engaged with the delivery catheter (i.e., “mapping” stage). With the LP at an acceptable location (PCT ≤ 3.0 V, sensed amplitude ≥ 1.0 mV), the delivery catheter control knob was used to rotate the LP until complete helix-myocardium engagement, as evident by 1.25-1.5 rotations of the radiopaque LP chevron (**Figure 2**B). In a minority of patients, the LP was moved to an alternative site and mapping was repeated.

The LP was then undocked from the delivery catheter docking cap, while still maintaining contact by the two delivery catheter tethers (i.e., “tethered” stage, **Figure 2**C), thus allowing mechanical and electrical evaluation, absent any substantial delivery catheter forces. LP mechanical stability was verified fluoroscopically by a catheter deflection test, and electrical contact was confirmed by repeat PCT, sensed amplitude, pacing impedance, and COI measurements.

With the LP engaged at an acceptable location, it was released from the tethers using the LP release knob, the catheter was removed (i.e., “released” stage, **Figure 2**D), and final acceptable electrical measurements were verified. Prior to patient discharge, the PCT, sensed amplitude, pacing impedance were measured. Any adverse events observed during the implant procedure or within 30 days post-implant were noted.

### Quantitative Analysis

For each LP implant procedure, the number of distinct mapping sites and deployment sites were noted, as was the final release location. Electrical metrics (i.e., PCT, sensed R-wave amplitude, and pacing impedance) were measured immediately upon LP release for all patients. In a subset of patients, electrical metrics were also measured with the LP still tethered, prior to release. Tether vs. release differences were evaluated using Wilcoxon signed-rank tests, with P<0.05 deemed significant.

The LP implant procedure duration was quantified as the time from delivery catheter introduction to removal. The fluoroscopy duration was quantified as the sum of all discrete uses of fluoroscopy related to LP implantation. Continuous variables are presented as median [interquartile range], including full range (min-max) when warranted. Adverse event-free rate and implant success rate are reported with Clopper-Pearson 95% binomial proportion confidence intervals.

## Results

### Study Population

A total of 167 patients were evaluated at 4 North American centers, with baseline patient characteristics provided in **Table 1**. Single-chamber pacemaker indications in this cohort predominantly included 3^rd^ degree AV block (38.3%), permanent atrial fibrillation (29.3%), tachy-brady syndrome (23.4%), sinus node dysfunction (19.8%), and paroxysmal atrial fibrillation (18.0%). Note that some patients presented with multiple indications.

**Table 1.**
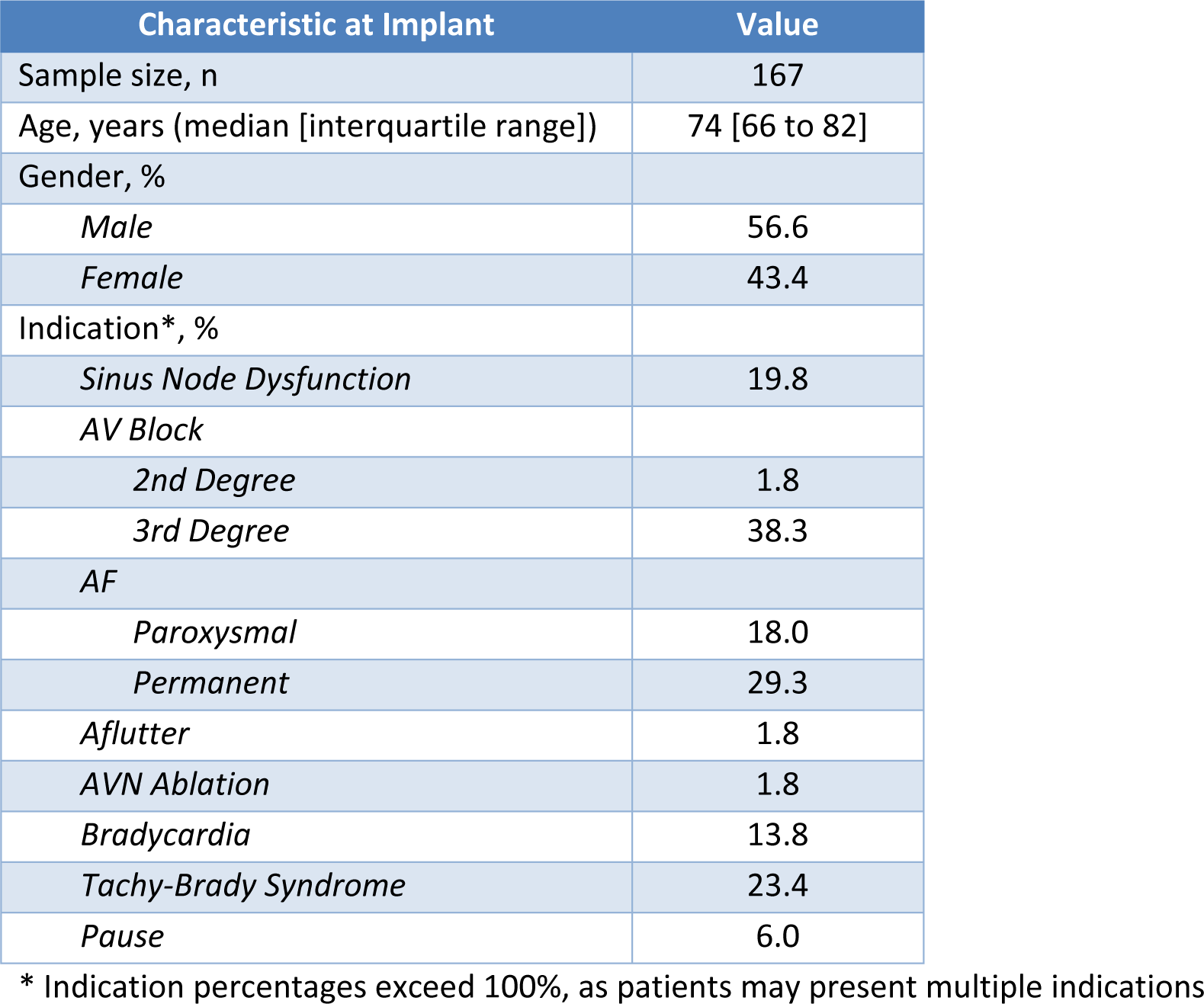
Baseline patient characteristics.

### Implant Procedure Characteristics

The implant procedure characteristics for this population are shown in **Figure 3**. The pre-fixation electrical mapping capability, unique to these helix-fixation LPs, resulted in one site mapped in 74.7% of patients, two sites mapped in 21.6% of patients, and 3-5 sites mapped in the remaining 3.7% of patients. This mapping capability ultimately resulted in the LP being deployed without post-fixation repositioning in 95.7% of patients; a single repositioning attempt was required in the remaining 4.3% of patients, including intraprocedural retrieval and reimplantation due to elevated PCT in 1.2% of patients.

**Figure 3.**
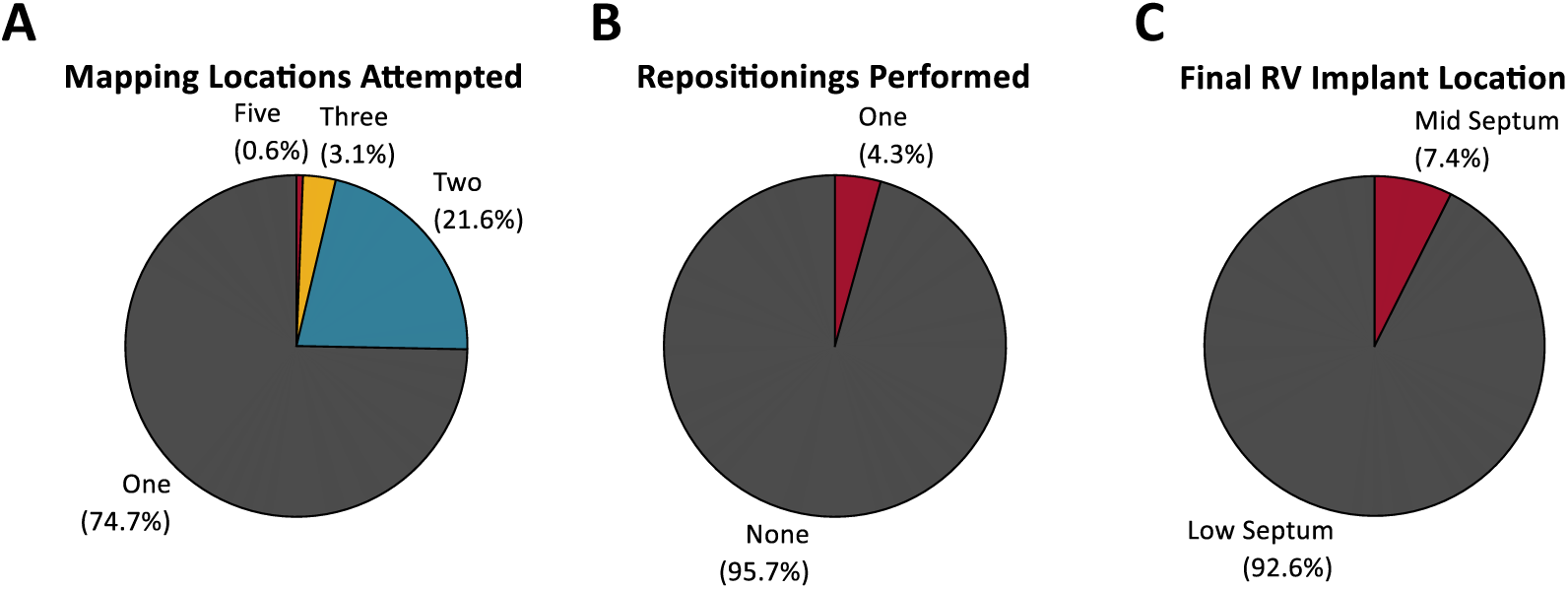
LP implant procedure population characteristics. Proportion of patients by (A) number of mapping locations attempted prior to implant, (B) number of device repositioning instances performed prior to release, and (C) final implant location.

LPs were predominantly implanted in the low RV septum (92.6%), with placement in the mid septum in the remainder. The median total procedure duration was 25.5 min [20.0, 35.0] (range: 14.0- 107.0), with a median fluoroscopy duration of 5.7 min [4.0, 9.2] (range: 1.4-44.9), as shown in **Figure 4**.

**Figure 4.**
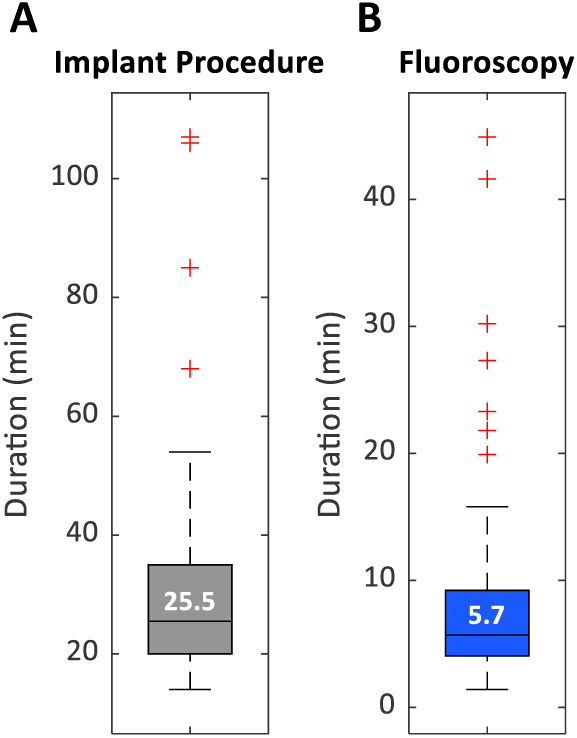
Implant procedure durations. (A) LP procedure duration, from delivery catheter introduction to retrieval, and (B) total cumulative fluoroscopy duration. Standard box plots show median (horizontal line), interquartile range (box), non-outlier range (dashed whiskers), and individual outlier values beyond 1.5x [interquartile range] from each of the two quartiles (red “+” symbol).

### Electrical Performance

Immediately after LP deployment and release, the electrical measurements were as follows (see **Table 2**). The median PCT at 0.4 ms pulse-width was 0.8 V [0.5, 1.3] (range: 0.3-4.5); sensed R-wave amplitude 9.0 mV [6.0, 12.0] (range: 2.0-28.0); pacing impedance 705 Ω [550, 910] (range: 310-1800). A PCT ≤ 2.0 V was observed in 96.3% of patients, and PCT ≤ 3.0 V were observed in 99.4% of patients (i.e., allowing a 2x safety margin with a maximum pulse amplitude of 6.0 V). Sensed R-wave amplitudes ≥ 3.0 mV (i.e., allowing sensing at the nominal VLP sensitivity threshold) were observed in 99.4% of patients, with amplitudes ≥ 1.0 mV observed in 100.0% of patients. While the current-of-injury electrograms were monitored, they were not quantitatively evaluated in this study.

**Table 2.** Aveir VR electrical measurements at implant, immediately after LP release.

| Electrical Measurement | Median [Interquartile Range] | Sample Size |
| --- | --- | --- |
| Pacing Capture Threshold @ 0.4 ms, V | 0.8 [0.5, 1.3] | 164 |
| Sensed R-wave Amplitude, mV | 9.0 [6.0, 12.0] | 156 |
| Pacing Impedance, $\Omega$ | 705 [550, 910] | 164 |

Changes in electrical measurements between the procedural state when the LP was still tethered to the delivery catheter (i.e., could still be re-docked and repositioned) and when the LP was subsequently released are shown in **Figure 5**. In the subset of patients for which pre-release “tethered” measurements were captured (N=31/167), median tether-release changes in PCT, sensed R-wave amplitude, and impedance of -0.5 V [-0.9, -0.3] (P<0.001), +0.5 mV [-1.0, 2.0] (P=0.205), and -30 Ω [-68, 25] (P=0.069) were observed.

**Figure 5.**
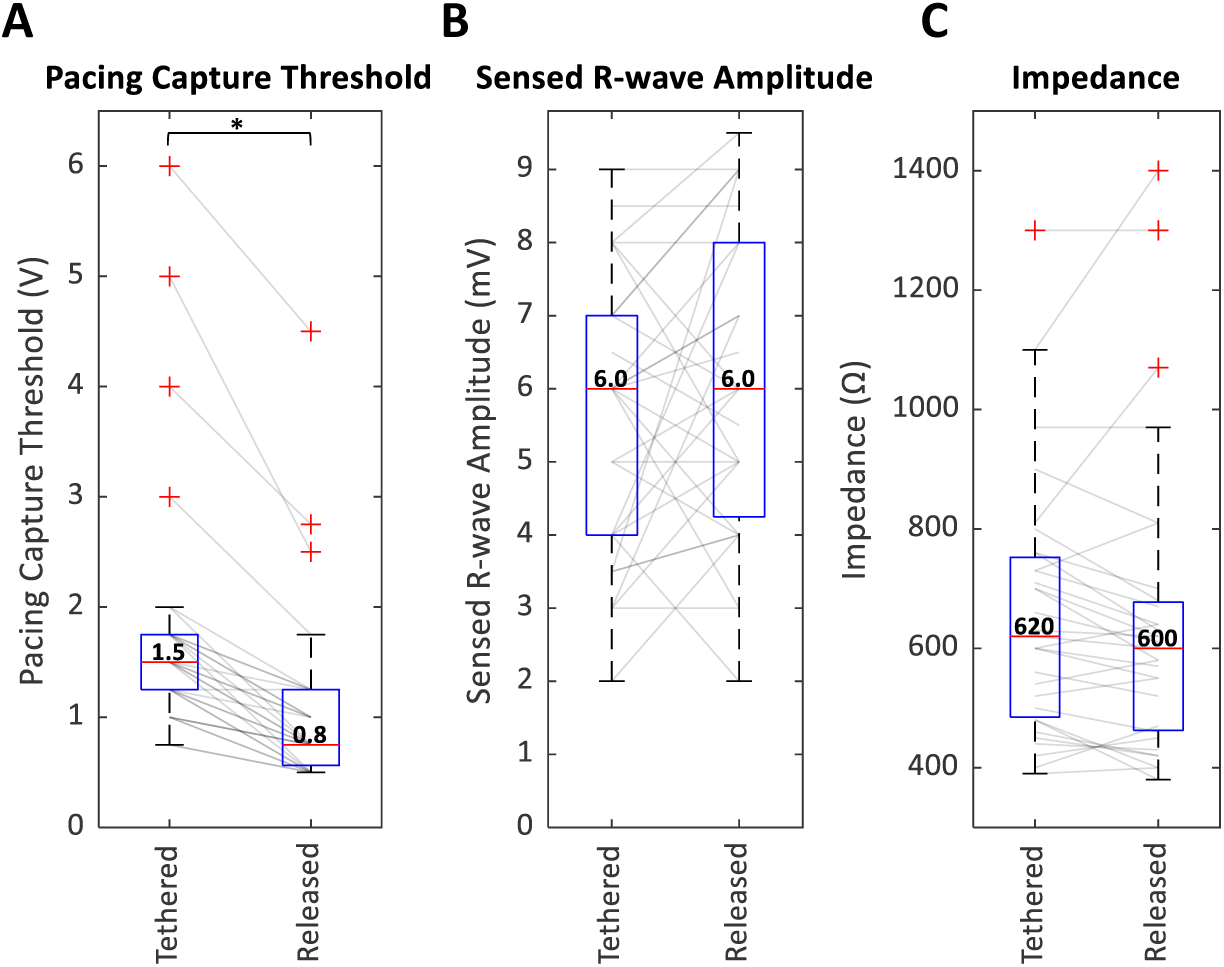
Changes in LP electrical measurements from when the device is tethered vs. released from the delivery catheter. (A) Pacing capture threshold at 0.4 ms, (B) sensed R-wave amplitude, and (C) pacing impedance are shown for the patient subgroup (N=31) with available data for both tether and release. Asterisk indicates P<0.05.

### Adverse Events

The distribution of 30-day adverse events related to the Aveir VR implant procedure are provided in **Figure 6**. Adverse events were absent in 98.2% of patients (95% confidence interval: 94.8- 99.6%), and were observed in three patients (1.8%). These include one patient with a pericardial effusion resulting in aborting of the implant procedure, sternotomy, stitch repair, and placement of epicardial leads (0.6%); one patient with delivery catheter tether fatigue during redocking before re-implantation with new device (0.6%); and one patient with leadless pacemaker dislodgement on the first post-procedural day, requiring device re-capture and replacement with a transvenous system (0.6%). Overall, the LP implants were completed successfully in 98.8% of patients (95% confidence interval: 95.7-99.9%), with two implants aborted due to the aforementioned pericardial effusion and dislodgement on the first post-procedural day.

**Figure 6.**
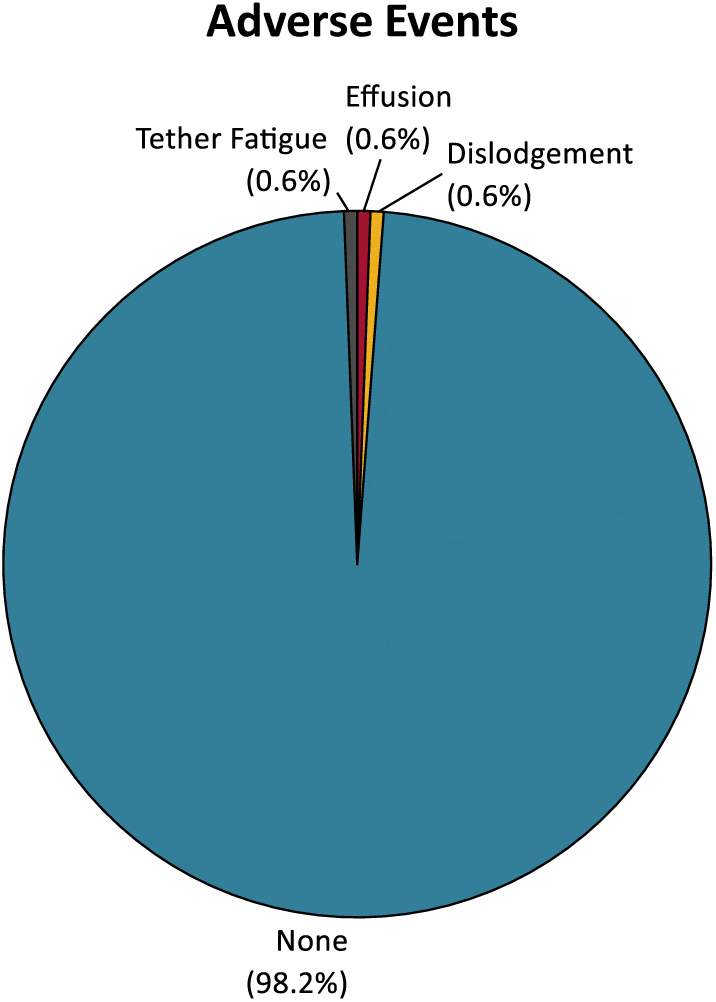
Distribution of adverse events related to Aveir VR implant procedure.

## Discussion

This initial, multi-center experience with the helix-fixation ventricular leadless pacemaker demonstrated safe and efficient real-world implantation. This LP system includes several features designed to facilitate implant site selection, streamline the implant procedure, and establish secure electrical contact. This evaluation demonstrated swift implant procedures, viable electrical metrics, and minimal acute complications.

The mapping capability, unique to these helix-fixation leadless devices, allows electrical contact with the myocardium to be evaluated before committing to an implant site. This pre-deployment verification resulted in viable electrical metrics to be achieved with minimal repositioning, which may have played a role in minimizing median LP procedure and fluoroscopy times to 25.5 min and 5.7 min, respectively. Although the first mapped RV site exhibited adequate electrical measurements in most patients, additional sites were mapped prior to fixation in roughly 1 in 4 patients. Consequently, LP repositioning after fixation, which can potentially lead to downstream clinical complications, was avoided in 95.7% of patients; only one repositioning was required in the remaining 4.3%. In other words, no LP implants required more than one repositioning attempt. In contrast, a commercial evaluation of the tines-fixation LP (Micra VR) reported that 22.7% of those implants required more than one repositioning attempt (i.e., 3 or more deployments).^8^

Successful electrical contact between each LP and the host myocardium was evident by post-release LP measurements conducted via the device programmer during the implant procedure. The LP exhibited a median PCT value of 0.8 V at 0.4 ms pulse-width (96.3% of devices presenting PCT ≤ 2.0 V), sensed R-wave amplitude of 9.0 mV, and impedance of 705 Ω. Taken together, these electrical metrics demonstrate LP-myocardial contact capable of providing effective pacing and sensing after device deployment. These commercial electrical measurements are consistent with the results from the first-in-human experience with this helix-fixation LP in the LEADLESS II IDE Study (Phase 2), which reported similar mean implant PCT values of 0.8 V (0.4 ms pulse-width), mean sensed R-wave amplitudes of 8.8 mV, and mean impedance values of 796 Ω.^9^

The step-wise LP implant procedure allows electrical measurements to be verified at sequential stages of commitment to a particular implant site. In a subset of devices in which pre-release (i.e., “tethered”) and post-release measurements were captured, a significant PCT improvement (0.5 V, approximately 33%) with relatively stable sensed amplitude and impedance values was observed upon release by the delivery catheter tethers. PCT improvement after release may be attributed to (a) time elapsing from the acute current of injury imposed by the active fixation mechanism, and/or (b) the release of tension maintained by the delivery catheter tethers. These dynamics highlight the need to monitor changes in PCT in the initial post-implant phase, prior to repositioning of the device due to a slightly elevated PCT.

The delivery catheter, in conjunction with the loading tool and introducer, resulted in complication-free implants in 98.2% of patients and successful implants in 98.8%. These rates are consistent with the 96.2% complication-free rate and 97.6% implant success rate reported in the LEADLESS II IDE Study (Phase 2).^9^

### Limitations

As this was an evaluation of the initial experience with helix-fixation LP implants at 4 centers, the main limitation of the study was the sample size. Larger, real-world, multi-center studies are needed for a more comprehensive assessment of the implant procedure across broader implanter experience levels, patient population demographics, and RV implant sites. The limited sample size also precluded any statistical conclusions surrounding complications, which were infrequent.

Data regarding implant stability and electrical measurements were limited to the acute, post-implant period. While commercial projections beyond this time-point would be speculative, promising long-term implant stability and electrical measurements have been shown in clinical trials.^9^

## Conclusions

The initial, real-world experience of the mapping-capable helix-fixation ventricular leadless pacemaker demonstrated safe and efficient implantation with minimal repositioning, viable electrical metrics, limited adverse events during implant or recovery, and a 98.8% implant success rate—all consistent with results from the LEADLESS II IDE Study.

## Data Availability

All data from corresponding study sites.

